# Supplemental oxygen costs and equipment utilization in a low-income population with ILD or COPD

**DOI:** 10.1101/2024.11.23.24317811

**Authors:** Kristopher P. Clark, Daniel J. Kass, Howard B. Degenholtz

## Abstract

**Rationale:** Supplemental oxygen is often prescribed to patients with interstitial lung disease (ILD) and chronic obstructive pulmonary disease (COPD). The specific costs of oxygen therapy in these diseases have not been fully described in patients living in the United States.

**Objectives:** Estimate Medicaid/Medicare costs for supplemental oxygen, characterize differences in oxygen equipment utilization, and identify factors impacting the costs of supplemental oxygen therapy in ILD and COPD.

**Methods:** We reviewed claims data for years 2016-2020 for Pennsylvania residents with ILD or COPD who were dually eligible for Medicaid/Medicare and enrolled in traditional fee-for-service Medicare. A generalized estimated equation (GEE) model was used to identify variables associated with annual oxygen costs.

**Measurements and Main Results:** A greater proportion of paid claims in ILD were for oxygen services (40.3% vs 22.0%) and for high flow oxygen (4.2% vs 2.2%), though ILD represented the minority of paid oxygen claims (5.2%). Oxygen cost approximately $65/month for both groups. Most oxygen claims (≥94%) were for stationary concentrators. Liquid devices and stationary gas were the least utilized equipment. Lower costs were associated with living in a competitive bidding area and with markers of advanced age and worse health status.

**Conclusions:** Oxygen utilization is greater in ILD but is only a small portion of total oxygen claims compared to COPD. Costs and equipment utilization were similar between groups. Liquid oxygen claims were rare even among subjects requiring high flow. Oxygen therapy is common in these diseases and likely represents a significant contribution to total healthcare costs.

## INTRODUCTION

In the United States, supplemental oxygen therapy is recommended for patients with chronic obstructive pulmonary disease (COPD) and interstitial lung disease (ILD) who have hypoxemia at rest or with exertion(1). These recommendations are based primarily on studies of long-term oxygen therapy (LTOT) which show improved mortality in patients with COPD(2,3). Oxygen therapy is common in both diseases(4–7) and is associated with increased healthcare costs and resource utilization(6,8–12). Healthcare spending on chronic respiratory diseases is estimated at over $100 billion per year(13,14); however, limited data exists on the costs associated with oxygen therapy in ILD and COPD, and little is known about how these patients utilize oxygen equipment or what factors contribute to oxygen costs in these diseases.

Supplemental oxygen is delivered from stationary or portable compressed gas tanks, liquid oxygen reservoirs/canisters, or oxygen concentrators. These devices differ in weight, ease of use, capacity to provide high oxygen flow delivery, and cost(15). Patients with ILD and COPD are often eligible for Medicare which provides coverage for durable medical equipment (DME) services including supplemental oxygen equipment and supplies. The Centers for Medicare and Medicaid Services (CMS) provides coverage for rented oxygen equipment/supplies including a 50% payment increase for equipment delivering >4 L/min(16,17). While the prescription of oxygen therapy is determined by a medical provider, the type of equipment a patient receives is often at the discretion of the locally contracted DME supplier. In 2011, CMS instituted a competitive bidding program (CBP) designed to lower costs for oxygen and other DME(18,19). While competitive bidding reduced DME spending(20–22), outside observers and patient advocacy groups contend that these changes also resulted in a decrease in DME suppliers and impaired patients’ access to supplemental oxygen services(23–25). This has prompted the drafting of the Supplemental Oxygen Access Reform (SOAR) Act which aims to improve oxygen access and payment structure(26).

We chose to study oxygen costs and utilization in patients who were dually eligible and receiving both Medicaid and Medicare fee-for-service (FFS) benefits as this represents vulnerable, low-income patients likely to have high disease burden. This also allowed us to focus on CMS policy regarding supplemental oxygen payments. We conducted this study of oxygen claims in dually eligible Pennsylvania residents with a diagnosis of ILD or COPD. Our objectives were to 1) estimate the costs for supplemental oxygen therapy, 2) characterize differences in utilization of oxygen delivery equipment, and 3) identify factors associated with increased costs of supplemental oxygen therapy in these diseases.

## METHODS

### Data Source

We reviewed Pennsylvania Medicare and Medicaid claims paid between January 1, 2016, and December 31, 2020. The data include beneficiary demographics and line-item DME claim payment amounts. Competitive bidding areas (CBA) were determined by matching beneficiary data to 2021 CBA zip codes(27) which were stable when compared to zip code data from earlier years (**Supplement 1**).

### Study Population

Subjects were dually eligible to receive both Medicaid and Medicare benefits for ≥1 month in a calendar year. Subjects enrolled in Medicare Advantage or Special Needs Plans were excluded since encounter data were not available. Subjects were required to have ≥2 outpatient or ≥1 inpatient claims associated with a diagnosis of ILD or COPD based on International Disease Classification (ICD-10) coding (**Supplement 2**). ILD was defined by ICD J84 series which represents fibrotic and interstitial diseases including idiopathic pulmonary fibrosis (IPF), the most common form of ILD. COPD was defined using a validated coding algorithm developed by the Chronic Condition Data Warehouse (CCW)(28). Subjects were excluded if they received nursing home care or were non-Pennsylvania residents (**Supplement 3**).

### Supplemental Oxygen Use

Subjects were identified as oxygen users if they had ≥1 paid claim in a calendar year associated with a Healthcare Common Procedure Coding System (HCPCS) code for oxygen equipment (**Supplement 4**). Claims were analyzed based on equipment type (compressed gas, liquid, or concentrator), whether equipment was stationary or portable, and low versus high flow delivery. We defined high flow as equipment coded for providing >4 L/min to align with CMS reimbursement policy(16).

### Comorbidities

Comorbid conditions expected to be associated with supplemental oxygen use (tobacco use, heart failure, pulmonary hypertension) were included in the model. To control for underlying health status, we constructed a count of 27 other comorbid conditions based on the validated set of algorithms (1999–2021) available from the CCW(28). See **Supplement 5** for definitions and prevalence rates of comorbidities.

### Statistical Analyses

The primary outcome was total Medicare/Medicaid payments for oxygen equipment. Chi-squared test at alpha=0.05 was used to compare median payment amounts. The total monthly payment was calculated for each beneficiary using the total annual spending by both Medicaid and Medicare divided by the number of months of eligibility that year. Annualized payments were estimated by top/bottom-coding total monthly payment at the 95th-percentile to eliminate outliers and multiplying by 12. Subcategories with fewer than 10 observations are redacted.

A generalized estimating equation (GEE) was estimated using the number of months of eligibility per year as an offset term. This approach was chosen to account for repeated observations of beneficiaries across multiple years. The model was adjusted for demographic and other covariates described above. Beneficiaries with both ILD and COPD diagnoses were included in the ILD cohort for all analyses. A sensitivity analysis was performed with these subjects included in the COPD cohort. All analyses were performed using StataSE version 18.

This study was determined to be exempt from human subjects research by the Institutional Review Board at the University of Pittsburgh.

## RESULTS

A total of 2,297 unique beneficiaries with ILD and 72,412 with COPD were included in this study. Baseline characteristics are reported using the first year each beneficiary appeared in the data (**Table 1**). Mean (SD) age was 67.6 (14.0) for ILD and 64.5 (12.4) for COPD. A majority of subjects in both cohorts were female and non-Hispanic white. A majority of subjects in both cohorts were residents of non-rural areas and/or resided in a CBA. A total of 32.0% of ILD and 25.2% of COPD beneficiaries were receiving home and community-based services. More than half of the beneficiaries in both cohorts had ≥4 comorbidities. In the first year of study inclusion, a greater proportion of beneficiaries with ILD compared to COPD had claims paid for any oxygen equipment (34.5% vs 18.4%) and high flow oxygen (1.7% vs 0.5%).

**Table 1.** Demographic and clinical characteristics of unique individuals at time of first year in the dataset(2016–2020)

|  | **ILD (n=2,297)** | **COPD (n=72,412)** |
| --- | --- | --- |
| Age, mean (SD) | 67.6 (14.0) | 64.5 (12.4) |
| Male, N (%) | 901 (39.2) | 29,574 (40.8) |
| Race/Ethnicity, N (%) |  |  |
| *Non-Hispanic White* | 1,392 (60.6) | 51,578 (71.2) |
| *Non-Hispanic Black* | 420 (18.3) | 13,226 (18.3) |
| *Hispanic* | 218 (9.5) | 3,673 (5.1) |
| *Other* | 267 (11.6) | 3,935 (5.4) |
| Home and community-based services | 734 (32.0) | 18,269 (25.2) |
| Months with an oxygen claim, median (IQR) | 11 (5-12) | 12 (6-12) |
| Non-metro resident, N (%) | 330 (14.4) | 13,999 (19.3) |
| Competitive bidding area resident, N (%) | 1,489 (64.8) | 42,546 (58.8) |
| Confounders of oxygen use, N (%) |  |  |
| *Current or prior tobacco use* | 600 (26.1) | 30,235 (41.8) |
| *Heart failure* | 987 (43.0) | 21,252 (29.4) |
| *Pulmonary hypertension* | 457 (19.9) | 5,526 (7.6) |
| Count of other comorbidities, N (%) |  |  |
| *0 to 1* | 164 (7.1) | 8,243 (11.4) |
| *2 to 3* | 383 (16.7) | 17,342 (24.0) |
| *4 to 5* | 576 (25.1) | 19,141 (26.4) |
| *6 to 7* | 580 (25.3) | 15,190 (21.0) |
| *8 to 9* | 390 (17.0) | 8,886 (12.3) |
| *10 or more* | 204 (8.9) | 3,610 (5.0) |
| Any Oxygen Claim, N (%) | 792 (34.5) | 13,300 (18.4) |
| High Flow (>4 L/min) Oxygen Claim, N (%) | 37 (1.7) | 329 (0.5) |

The 2,297 ILD subjects had a total of 4,344 person-years during the study period (i.e., unique beneficiary who met the inclusion criteria could appear in more than one calendar year); 40.3% of person-years (1,752) had at least one claim for oxygen services. The 72,412 COPD subjects had a total of 144,816 person-years during the study period; 22.0% of person-years (31,864) had at least one claim for oxygen services. ILD subjects had a greater proportion of oxygen claims across all five study years (**Supplement 6**). Of all person-year claims for oxygen services, a greater proportion of ILD claims were coded high flow oxygen (4.2%) compared to COPD (2.2%). Despite having a greater proportion of oxygen claims, ILD represented only 5.2% of all oxygen claims and 9.0% of all high flow claims.

The total amount of oxygen payments made during the five-year study period was $22,216,939 of which 76.5% ($16,994,808) were made by Medicare. Of the total combined Medicare/Medicaid payment amount during the study period, 5% ($1,107,372) was for ILD and 95% ($21,109,567) was for COPD. The median (IQR) monthly payment for all oxygen claims was $64.63 ($35.04-$94.56) for ILD and $65.84 ($33.63-$96.70) for COPD (p-value=0.985). Median monthly claim payments by calendar year ranged from $59.57-$74.11 for ILD and $56.57-$78.39 for COPD (**Figure 1**).

**Figure 1.**
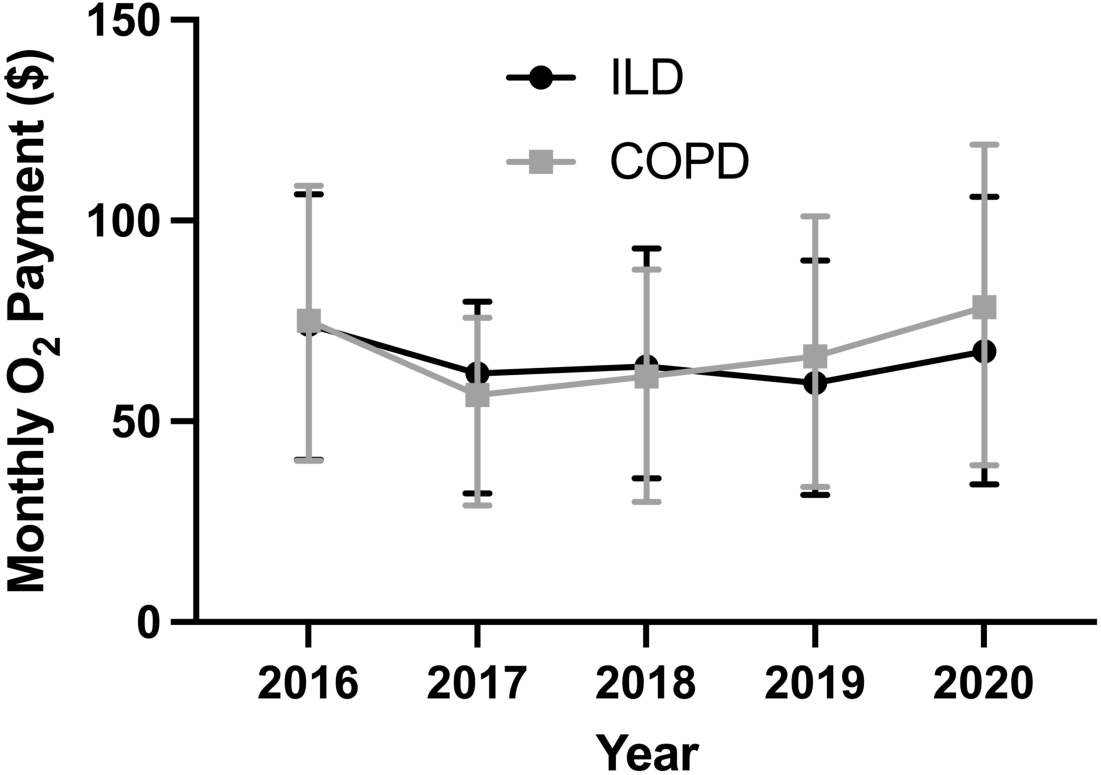
Median (IQR) monthly payment for oxygen claims by year.

At least 94% of paid oxygen claims in both cohorts included stationary oxygen concentrators. A total of 69.2% of ILD and 62.8% of COPD claims included payments for portable gas cannisters while 16.3% of ILD and 11.7% of COPD claims included payments for portable concentrators. Less than 2.5% of claims in both cohorts included liquid oxygen (portable or stationary). A similar utilization pattern was seen for claims coded for high flow delivery (**Figure 2**).

**Figure 2.**
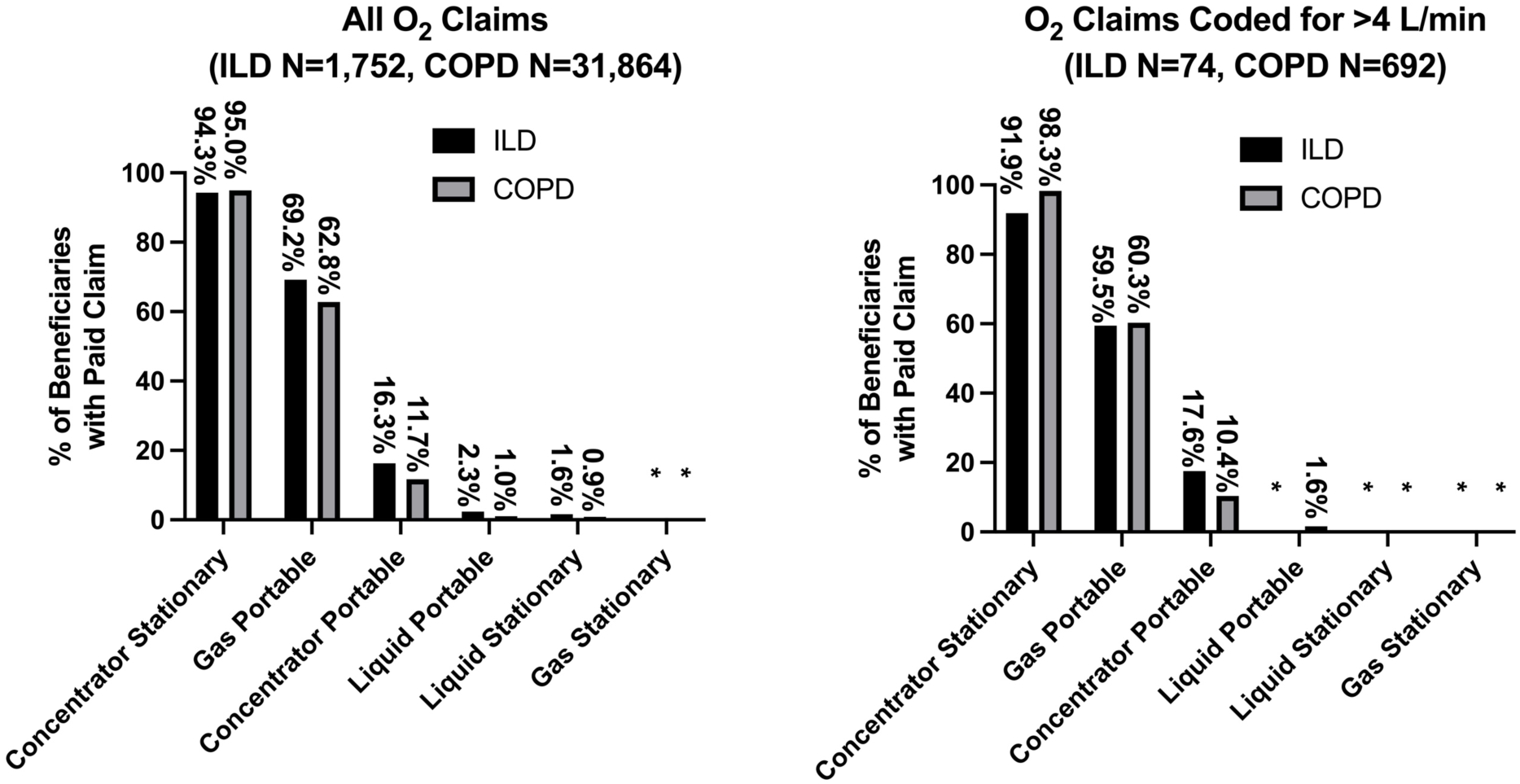
Proportion of person-years claims by equipment type. **Note: Categories with <10 observations are redacted**

Median monthly payments varied by equipment type and ranged from $12.92-$54.67 for ILD and $13.78-$56.83 for COPD. For all paid claims and for high flow claims, the highest payment was for stationary concentrators and the lowest payment was for portable gas. With the exception of portable gas (p=0.020), there was no significant difference in median monthly equipment payments between cohorts. In comparing payments for low vs high flow equipment, high flow stationary concentrator payments were greater for COPD only (p-value <0.001). All other equipment payments for high flow claims were not significantly different from low flow claims (**Table 2**)

**Table 2.** Median (IQR) monthly payment for person-year claims by oxygen equipment type and flow rate.

| Equipment Type | ILD |  |  |  | COPD |  |  |  |
| --- | --- | --- | --- | --- | --- | --- | --- | --- |
|  | All claims | >4 L/min claims | ≤4 L/min claims | p-value* | All claims | >4 L/min claims | ≤4 L/min claims | p-value* |
| Stationary concentrator | \$54.67<br>(\$29.62-\$74.27) | \$55.03<br>(\$30.20-\$72.03) | \$54.67<br>(\$29.55-\$74.53) | 0.597 | \$56.83<br>(\$29.55-\$80.00) | \$61.61<br>(\$35.89-\$86.16) | \$56.74<br>(\$29.19-\$79.78) | <0.001 |
| Portable concentrator | \$21.20<br>(\$11.72-\$28.96) | \$22.21<br>(\$12.88-\$25.32) | \$21.08<br>(\$11.35-\$29.19) | 0.806 | \$22.77<br>(\$11.41-\$29.01) | \$19.37<br>(\$9.54-\$29.99) | \$22.79<br>(\$11.42-\$29.01) | 0.28 |
| Portable gas | \$12.92<br>(\$6.64-\$21.92) | \$12.56<br>(\$6.60-\$21.20) | \$12.96<br>(\$6.65-\$22.01) | 0.113 | \$13.78<br>(\$7.53-\$22.38) | \$14.02<br>(\$8.58-\$23.57) | \$13.78<br>(\$7.51-\$22.36) | 0.452 |
| Portable liquid | \$16.62<br>(\$7.77-\$31.27) | -- | \$20.18<br>(\$9.90-\$31.69) | -- | \$18.83<br>(\$11.59-\$32.96) | \$19.28<br>(\$10.13-\$29.51) | \$18.82<br>(\$11.59-\$32.99) | 0.143 |
| Stationary gas | -- | -- | -- | -- | -- | -- | -- | -- |
| Stationary liquid | \$43.05<br>(\$29.52-\$59.99) | -- | \$37.33<br>(\$24.63-\$59.75) | -- | \$45.22<br>(\$30.72-\$69.02) | -- | \$45.22<br>(\$31.16-\$68.73) | -- |

\*p-value compares median payment between >4 L/min and ≤4 L/min claims
Note: cells with <10 observations are redacted

A GEE model for repeated measures was performed to identify variables associated with higher annual oxygen payments among those with paid oxygen claims. As nearly all claims (≥94%) in both cohorts included stationary oxygen concentrators, we restricted the analysis to only these subjects and defined the dependent variable as the additional cost above and beyond that for a stationary concentrator. The model was adjusted for age, sex, race, residence in a non-metro area or CBA, long-term services and support use, diagnoses that may impact oxygen use (tobacco use, heart failure, pulmonary hypertension), count of other comorbidities, whether the oxygen claim was coded for high flow, and primary diagnosis (ILD vs COPD). Due to small sample sizes, we combined stationary and portable services for gas and liquid oxygen (e.g., gas stationary or portable, liquid stationary or portable).

**Table 3a** shows the results of the GEE model. Of beneficiaries who had payments for stationary concentrators, variables significantly associated with higher annual oxygen claim payments (p-value <0.05) included race other than Hispanic or non-Hispanic white or black, living in a non-metro location, having a diagnosis of heart failure, receiving gas or liquid oxygen or receiving portable concentrators in addition to a stationary concentrator. Increasing age, receiving home-based care, residing in a CBA, current or prior tobacco use, and increasing number of comorbidities were associated with lower annual oxygen claim payments. Increasing age and number of comorbidities were inversely related to payments for additional ambulatory oxygen equipment. Sex, a diagnosis of pulmonary hypertension, whether the oxygen claim was coded for high flow, and a diagnosis of ILD were not significantly associated with annual oxygen claim payments. The cost of oxygen varied based on the combination of equipment that subjects receive. Subjects who only received stationary concentrators had estimated annual costs of $964 for ILD and $945 for COPD. Subjects who had both stationary concentrators and liquid oxygen had an estimated annual cost $1,164 for ILD and $1,144 for COPD. The estimated annual cost for subjects with a stationary oxygen concentrator and gas oxygen was $1,027 for ILD and $1,008 for COPD. The estimated annual cost for subjects with both stationary and portable oxygen concentrators was $1,241 for ILD and $1,222 for COPD (**Table 3b**).

**Table 3.**
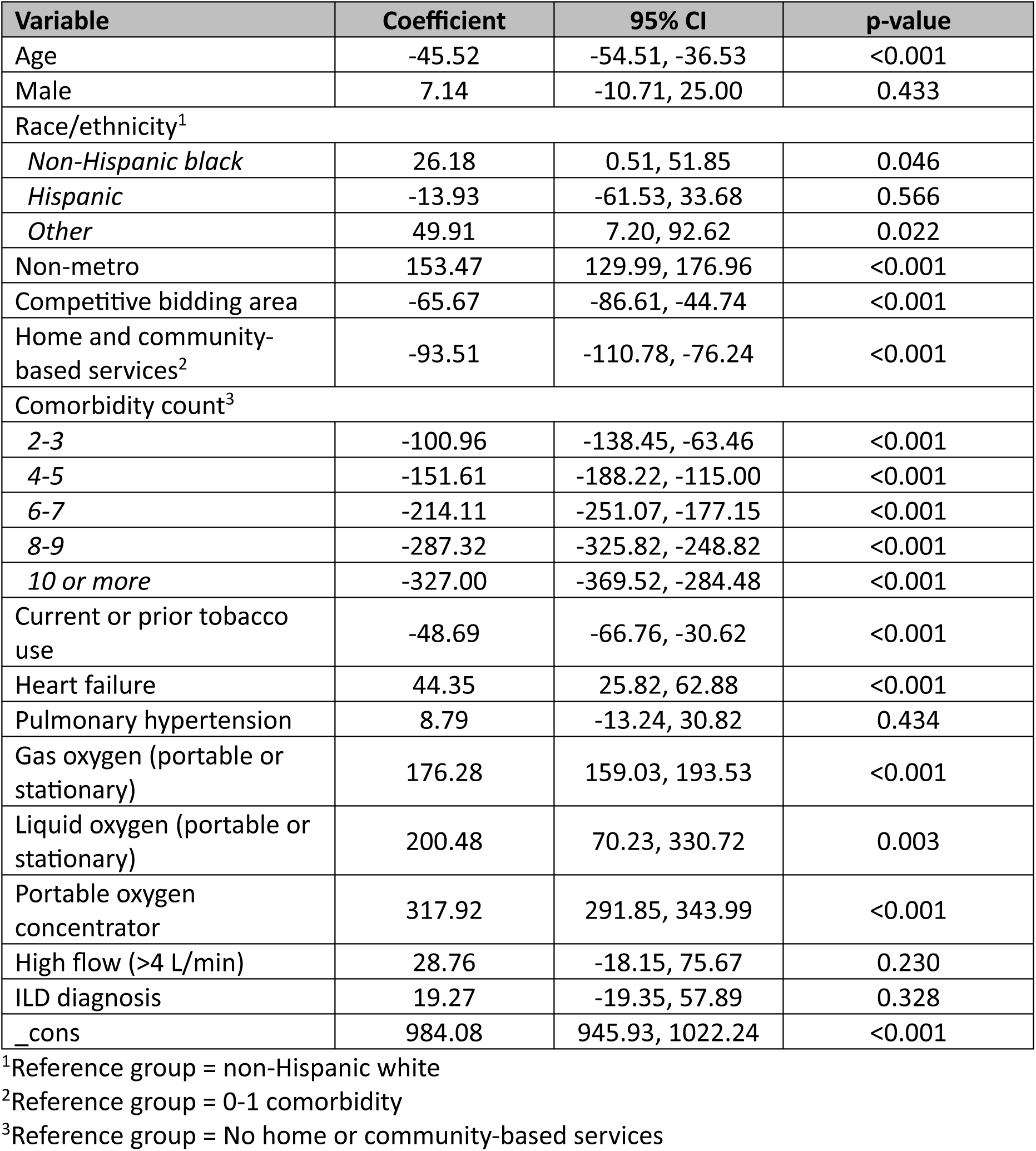
Results of GEE model. Table 3a. Results of a GEE model for repeated measures assessing factors influencing oxygen payments among beneficiaries who had claims paid for stationary oxygen concentrators

**Table 3b.**
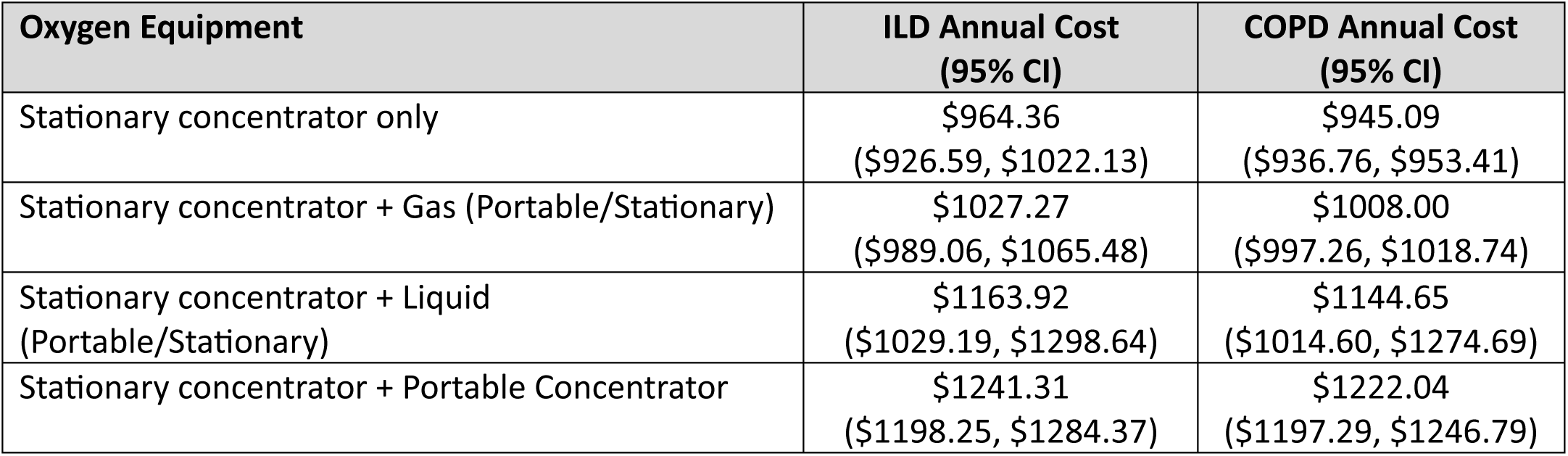
Estimated annual costs of oxygen equipment based on GEE model.

In a sensitivity analysis where beneficiaries with dual ILD and COPD diagnoses were included in the COPD cohort, baseline characteristics and findings from the final GEE model were similar with the exception being that there was no longer a significant difference in the number of oxygen claims at baseline between cohorts (p-value=0.754) (**Supplement 7**).

## DISCUSSION

The results of our study demonstrate that among Pennsylvania residents dually eligible to receive Medicare and Medicaid, a greater proportion of beneficiaries with ILD compared to COPD used supplemental oxygen and were more likely to have a high flow rate (>4 L/min); however, COPD beneficiaries represented the vast majority (95%) of all paid oxygen claims. The utilization of equipment other than oxygen concentrators was low in both groups even among those receiving high flow rates. Having additional oxygen equipment was associated with increased cost but was less common among older people and those with multiple chronic conditions.

ILD and COPD are associated with high health care costs and health resource utilization(6,8–12). In a study examining health care spending between 1996 and 2016, both ILD and COPD were identified as being among the top 100 most expensive health conditions with spending in 2016 estimated at $2.1 billion and $34.3 billion respectively(13). While these estimates did not include DME costs, other studies have found that DME use and specifically the use of supplemental oxygen are also associated with increased healthcare costs and utilization(6,11,12). Physician/DME services contribute an additional $100 billion in Medicare/Medicaid expenses each year(29). Our findings add to the existing literature by providing estimates on the cost of oxygen services in ILD and COPD in low-income patients living in the U.S.

We also found that payments for specific oxygen delivery systems did not vary significantly between ILD and COPD beneficiaries. We found that the vast majority of paid oxygen claims (≥94%) included stationary concentrators and that stationary concentrators were associated with the highest median monthly payment at approximately $55-$57. Our findings align with those of a prior study of CMS DME claims from 2013-2019 which identified stationary oxygen concentrators as the highest volume respiratory DME product, utilized by 2.32% of all Medicare beneficiaries in 2019(30). Both this study and our study identified low utilization rates for other delivery devices including liquid oxygen. Society guidelines recommend portable liquid oxygen for patients requiring >3 L/min and who are active outside the home(1); however, we found that only 2.9% of all high flow portable oxygen claims were for portable liquid oxygen. It is possible that liquid oxygen utilization rates are low due to decreased availability of this device, potentially as a result of CMS’s competitive bidding program (CBP). Our estimated monthly median costs differ from those found in a recent review of supplemental oxygen spending in Medicare beneficiaries with COPD which estimated monthly mean costs ranging between $687 to $771(22). This may be due to different methodologies in cost estimation and which payments were included. Our study focused only on payments made by Medicaid/Medicare and did not include other insurance payments or beneficiary out-of-pocket expenses.

As previously noted, CMS implemented the CBP in 2011 as a measure to decrease CMS expenditures and beneficiary out-of-pocket expenses on selected DME with high utilization rates and high costs(18). Oxygen equipment is one of the most frequently prescribed DME and is included in the CBP(31). Our model found that living in a CBA was associated with lower oxygen costs, consistent with reports by CMS and other government agencies that the CBP lowered costs of DME(31–33). These agencies also report that the CBP has not negatively impacted clinical outcomes or disrupted DME supply, assertions that were also noted in a recent review of early CBP oxygen claims data in patients with COPD(22). However, numerous outside observers including researchers and advocacy groups have raised concerns that the CBP has resulted in a decreased number of DME suppliers and disrupted access to DME services including liquid oxygen(23–25,34). It has been reported that the number of DME suppliers contracted by CMS to provide oxygen equipment has decreased over time and that the percentage of beneficiaries using oxygen DME has also decreased with the greatest change seen with stationary (−89%) and portable (−86%) liquid oxygen devices(30). The reduction in liquid oxygen utilization has raised concerns that the CBP resulted in more limited access to this therapy(23–25,35). Liquid oxygen is considered to be a more ideal delivery method for patients who require high flow rates due to its lighter weight and longer duration of use with higher flows compared to compressed gas tanks; however, it is likely more costly for DME suppliers to maintain and deliver this form of oxygen therapy which may limit its availability. While many stationary concentrators can meet high flow demands, portable oxygen concentrators currently can not. This limits the portable oxygen options for patients requiring higher flow. The reduced availability and utilization of portable liquid oxygen may have negative impacts on patients’ mobility and quality of life. These concerns have led to the drafting of the Supplemental Oxygen Access Reform (SOAR) Act which aims to improve oxygen access and change how oxygen equipment is reimbursed(26). While we did not compare oxygen costs or utilization over time or to pre-competitive bidding data, our findings, which included both COPD and ILD diagnoses, suggest that there are limitations in oxygen equipment utilization, at least among low-income patients including those who require high flow oxygen. It is possible these limitations resulted from changes implemented by the CBP.

In addition to living in a CBA, we also found that advanced age and increased number of comorbidities were associated with reduced oxygen payments. Further analysis revealed that these findings were primarily related to reduced payment for portable oxygen devices. Whether this means that older and sicker patients are less mobile or less likely to undergo ambulatory oxygen assessments is not known. Lastly, current or prior tobacco use was associated with a lower cost, possibly due to prescribers withholding oxygen prescriptions to patients who actively smoke due to increased risk of fire and injury.

Increased oxygen costs were associated with living in a non-metro area. Greater than 98% of non-metro claims are excluded from a CBA. We also found that non-Hispanic black and non-Hispanic, non-white or non-black race were associated with higher costs compared to non-Hispanic whites. Both of these race/ethnic groups had a greater proportion of beneficiaries residing within CBAs compared non-Hispanic whites. Racial and ethnic minorities have been shown to bear a disproportionate economic burden related to healthcare costs secondary to inequities in disease burden, morbidity, and premature death(36). Such inequities may also be found in oxygen equipment utilization as well which may account for this finding. Future studies should further explore the impact race/ethnicity, income, and education have on oxygen utilization.

A diagnosis of heart failure was also associated with increased costs while pulmonary hypertension was not. Pulmonary hypertension is often associated with oxygen needs and may be more common in patients with ILD(37,38). It is possible that our study was underpowered to detect the true effect of this comorbidity. Lastly, while CMS provides 50% increased payment to claims coded for providing high flow (>4 L/min)(17), with the exception of stationary concentrators for COPD, we did not find that payments for high flow claims were significantly greater than claims for ≤4 L/min. It may be that our study was underpowered to detect such differences given the low number of high flow claims in our dataset.

Our study has several strengths. To our knowledge, we are the first study to quantify the cost of oxygen spending in ILD in the U.S. and compare these costs to COPD. We were able to maximize our sample size by using claims data across five years which allowed us to not only quantify the use and cost of oxygen but to assess the impact other parameters have on oxygen costs. By focusing the study on dually eligible beneficiaries, we were able to estimate actual Medicare and Medicaid expenses for oxygen supplies for a vulnerable and policy-relevant population.

Our study also has several limitations. Our results may not be generalizable to beneficiaries receiving other Medicare or Medicaid services, those with substantial out-of-pocket expenses, or those who have oxygen services paid through private insurance carriers. Results may not be generalizable to patients living in other states; however, Pennsylvania is a large state with both large metropolitan areas and large rural areas and is demographically similar to the US average. Nevertheless, recent data from our center suggest that regional differences in air quality may adversely impact the prognosis of pulmonary fibrosis(39), thus oxygen utilization may have regional variation due to these exacerbating factors. Due to increased enrollment in Medicare Advantage plans over the course of our study, our data had a decreasing number of dually eligible individuals over time. We also did not calculate how oxygen costs compare to total DME expenditures or total Medicare and Medicaid spending. Future studies should examine the cost of oxygen in other populations including privately insured individuals and in national claims databases.

While our study identified payments made for oxygen claims, we were unable to determine whether oxygen prescriptions were appropriate or whether beneficiaries utilized these oxygen services. Lastly, our small sample of beneficiaries with high flow claims, especially among subjects with ILD, limits our ability to estimate oxygen costs and utilization for these subjects, though it does appear that, for some oxygen equipment, payments are not being increased as per CMS policy. Future studies and investigations are needed to further explore this finding.

## CONCLUSION

The vast majority of supplemental oxygen claims for Pennsylvania residents dually eligible for Medicaid and Medicare fee-for-service benefits with diagnoses of ILD or COPD were for stationary oxygen concentrators. Subjects with ILD had a greater proportion of paid oxygen claims but represent only a small portion of all oxygen claims. Among users of oxygen concentrators, the addition of liquid oxygen further increases costs compared to gas equipment, however the utilization of liquid oxygen is low in both diseases. Our findings support the concerns raised by other studies that the CMS competitive bidding program may have resulted in decreased access for liquid oxygen services.

## Data Availability

Data used in this study cannot be made available due to agreements with the Commonwealth of Pennsylvania.

## ACKNOWLEDGEMENTS

Jie Lin, PhD

