## Supplement for "Supplemental oxygen costs and equipment utilization in a low-income population with ILD or COPD"

### Supplement 1 – Zip Code Data Used to Determine Competitive Bidding Areas (CBA)

Beneficiaries were determined to reside in a CBA if their zip code was included in the Centers for Medicare and Medicaid (CMS) 2021 CBA Zip Code File: https://www.dmecompetitivebid.com/cbic/cbicr2021.nsf/DocsCat/H5O2KFK4HO.

We compared the 2021 CBA zip code file to the zip code files for the following CBA periods and determined that CBA zip codes remained relatively stable during the study period:

- Round Recompete: https://www.dmecompetitivebid.com/palmetto/cbicrd1recompete.nsf/DocsCat/Competitive%20Bidding%20Areas
- Round 2: https://www.dmecompetitivebid.com/palmetto/cbicrd2.nsf/DocsCat/Competitive%20Bidding%20Areas
- Round 2 Recompete: https://www.dmecompetitivebid.com/palmetto/cbicrd2recompete.nsf/DocsCat/Home
- Round 1 2017: https://dmecompetitivebid.com/palmetto/cbicrd12017.nsf/DocsCat/Competitive%20Bidding%20Areas

We identified 3 zip codes included in the 2021 CBA file that did not appear to be part of a CBA in CY2016 (16242, 18011, 18216). A total of 26 records in CY2016 were from these 3 zip codes. We conducted our analysis of baseline data and the GEE model with these zip codes labeled as CBA and non-CBA and did not find any significant differences. For simplicity, these zip codes are included as CBAs in the data presented in this manuscript.

**Supplement 2 – ICD-10 Diagnoses Codes for Interstitial Lung Disease (ILD) and Chronic Obstructive Pulmonary Disease (COPD) cohorts**

**Subjects were included if they had 2 outpatient or 1 inpatient encounter(s) with a primary diagnosis code as follows:**

**ILD:** J84.1, J84.10, J84.11, J84.111, J84.112, J84.113, J84.114, J84.115, J84.116, J84.117, J84.17, J84.170, J84.178

**COPD*:** J.40, J41.0, J41.1, J41.8, J42, J43.0, J43.1, J43.2, J43.9, J44.0, J44.1, J44.9, J47.0, J47.1, J47.9, J98.2, J98.3
*as per validated algorithm developed by the Chronic Condition Data Warehouse (https://www2.ccwdata.org)

### Supplement 3 – Selection process by calendar year

### 2016

**234,437** Dually eligible FFS beneficiaries

**203,110** Excluded
- 45,604 nursing home care
- 156,891 not COPD or ILD
- 165 non-PA resident
- 450 $O_2$ claim not paid

**31,867** included in analysis

### 2017

**233,026** Dually eligible FFS beneficiaries

**201,554** Excluded
- 42,960 nursing home care
- 158,019 not COPD or ILD
- 118 non-PA residents
- 457 $O_2$ claim not paid

**31,472** included in analysis

### 2018

**232,752** Dually eligible FFS beneficiaries

**201,552** Excluded
- 41,780 nursing home care
- 159,111 not COPD or ILD
- 146 non-PA residents
- 515 $O_2$ claim not paid

**31,201** included in analysis

### 2019

**220,182** Dually eligible FFS beneficiaries

**190.371** Excluded
- 38,307 nursing home care
- 151,542 not COPD or ILD
- 87 non-PA residents
- 435 $O_2$ claim not paid

**29,811** included in analysis

### 2020

**196,143** Dually eligible FFS beneficiaries

**171,334** Excluded
- 35,491 nursing home care
- 160,652 non-COPD or ILD
- 46 non-PA residents
- 291 $O_2$ claim not paid

**24,809** included in analysis

**Supplement 4 – Healthcare Common Procedure Coding System (HCPCS) Supplemental Oxygen Codes**

***Definition of HCPCS oxygen codes included in our study:***

OXYGEN DEVICES

- E0424 – Stationary compressed gaseous oxygen system, rental; includes container, contents, regulator, flowmeter, humidifier, nebulizer, cannula or mask, and tubing
- E0431 – Portable gaseous oxygen system, rental: includes portable container, regulator, flowmeter, humidifier, cannula or mask, and tubing
- E0433 – Portable liquid oxygen system, rental; home liquefier used to fill portable liquid oxygen containers, includes portable containers, regulator, flowmeter, humidifier, cannula or mask, and tubing, with or without supply reservoir and contents gauge
- E0434 – Portable liquid oxygen system, rental; includes portable container, supply reservoir, humidifier, flowmeter, refill adaptor, contents gauge, cannula or mask, and tubing
- E0439 – Stationary liquid oxygen system, rental; includes container, contents, regulator, flowmeter, humidifier, nebulizer, cannula or mask, and tubing
- E1390 – Oxygen concentrator, single delivery port, capable of delivering 85% or greater oxygen concentration at the prescribed flow rate, each
- E1391 – Oxygen concentrator, dual delivery port, capable of delivering 85% or greater oxygen concentration at the prescribed flow rate, each
- E1392 – Portable oxygen concentrator, rental
- K0738 – Portable gaseous oxygen system, rental; home compressor used to fill portable oxygen cylinders; includes portable containers, regulator, flowmeter, humidifier, cannula or mask, and tubing

OXYGEN CONTENT

- E0441 – Stationary oxygen contents, gaseous, 1 month's supply = 1 unit
- E0442 – Stationary oxygen contents, liquid, 1 month's supply = 1 unit
- E0443 – Portable oxygen contents, gaseous, 1 month's supply = 1 unit
- E0444 – Portable oxygen contents, liquid, 1 month's supply = 1 unit
- E0447 – Portable oxygen contents, liquid, 1 month's supply = 1 unit, prescribed amount at rest or nighttime exceeds 4 liters per minute (LPM)

***Modifiers that indicate claim for equipment providing >4 L/min***

- QB
- QF
- QG
- QR

***HCPCS code by equipment category***

**GAS OXYGEN**

- ***Stationary:*** E0424, E0441

- ***Portable:*** E0431, K0738, E0443

**LIQUID OXYGEN**

- ***Stationary:*** E0439, E0442
- ***Portable:*** E0433, E0434, E0444, E0447

**OXYGEN CONCENTRATOR**

- ***Stationary:*** E1390, E1391
- ***Portable:*** E1392

**Supplement 5 – Definitions and prevalence of comorbidities**

With the exception of pulmonary hypertension and tobacco use, all comorbidities (including heart failure) were identified using validated coding algorithms for 27 comorbidities (1999-2021) developed by the Chronic Condition Data Warehouse (CCW) (https://www2.ccwdata.org/web/guest/condition-categories-chronic). We combined CCW comorbidities related to dementia, cancer, and serious mental illness into single variables as noted in footnotes below. Tobacco Use was identified from a validated CCW algorithm listed under Other Chronic Health Conditions (https://www2.ccwdata.org/web/guest/condition-categories-other).Pulmonary hypertension was defined as $\geq$1 inpatient or $\geq$2 outpatient claims for ICD-10 code series I27.

**Prevalence rates of comorbidities at time of beneficiaries' first appearance in the dataset (2016-2020):**

| Condition | ILD (N=2,297) | COPD (N=72,412) | p-value |
|---|---|---|---|
| Acute myocardial infarction | 113 (4.9%) | 2,277 (3.1%) | <0.001 |
| Anemia | 1,301 (56.6%) | 29,730 (41.1%) | <0.001 |
| Asthma | 487 (21.2%) | 15,038 (20.8%) | 0.614 |
| Atrial fibrillation | 339 (14.8%) | 7,952 (11.0%) | <0.001 |
| Cancer[1] | 323 (14.06) | 7,513 (10.38) | <0.001 |
| Cataract | 262 (11.4%) | 6,978 (9.6%) | 0.005 |
| Chronic kidney disease | 1,093 (47.6%) | 26,261 (36.3%) | <0.001 |
| Dementia[2] | 379 (16.5%) | 8,472 (11.7%) | <0.001 |
| Diabetes | 944 (41.1%) | 27,738 (38.3%) | 0.007 |
| Glaucoma | 157 (6.8%) | 4,106 (5.7%) | 0.018 |
| Heart failure* | 987 (43.0%) | 21,252 (29.4%) | <0.001 |
| Hip fracture | 47 (2.1%) | 849 (1.2%) | <0.001 |
| Hyperlipidemia | 1,316 (57.3%) | 39,417 (54.4%) | 0.007 |
| Hypertension | 1,794 (78.1%) | 53,471 (73.8%) | <0.001 |
| Hypothyroidism | 497 (21.6%) | 12,512 (17.3%) | <0.001 |
| Ischemic heart disease | 1,196 (52.1%) | 29,265 (40.4%) | <0.001 |
| Osteoporosis | 310 (13.5%) | 5,014 (6.9%) | <0.001 |
| Prostatic hypertrophy | 215 (9.4%) | 5,268 (7.3%) | <0.001 |
| Pulmonary hypertension*[3] | 457 (19.9%) | 5,526 (7.6%) | <0.001 |
| Rheumatoid arthritis | 821 (35.7%) | 23,711 (32.7%) | 0.003 |
| Serious mental illness[4] | 1,088 (47.37) | 34,664 (47.87) | 0.634 |
| Stroke | 241 (10.5%) | 6,409 (8.9%) | 0.007 |
| Tobacco* | 600 (26.1%) | 30,235 (41.8%) | <0.001 |

Note: cells with <10 observations are redacted

[1]Cancer combines CCW data for breast, colorectal, endometrial, lung, and prostate cancers

[2]Dementia combines CCW data for Alzheimer's and other dementias

[3]Pulmonary hypertension is not included in the CCW but was defined as having $\geq$1 inpatient or $\geq$2 outpatient claims for ICD-10 code series I27

[4]Serious mental illness combines CCW data for anxiety, bipolar, and depressive disorders

[5]Tobacco was defined by CCW algorithm outlined under Other Chronic Health Conditions (https://www2.ccwdata.org/web/guest/condition-categories-other)

*Tobacco, heart failure, and pulmonary hypertension are included separately in our analysis, all other comorbidities are included in comorbidity counts

**Supplement 6 – Total number of beneficiaries and number of beneficiaries with an oxygen ($O_2$) claim by claim year**

| Year | ILD | | COPD | |
|---|---|---|---|---|
| | Beneficiaries | With Any $O_2$ Claims, N (%) | Beneficiaries | With Any $O_2$ Claim, N (%) |
| 2016 | 866 | 381 (44.0%) | 31,001 | 7,205 (23.2%) |
| 2017 | 857 | 356 (41.5%) | 30,615 | 6,845 (22.4%) |
| 2018 | 933 | 375 (40.2%) | 30,268 | 6,538 (21.6%) |
| 2019 | 895 | 350 (39.1%) | 28,916 | 5,939 (20.5%) |
| 2020 | 793 | 290 (36.6%) | 24,016 | 5,337 (22.2%) |
| **Total** | **4,344** | **1,752 (40.3%)** | **144,816** | **31,864 (22.0%)** |

**Supplement 7 – Sensitivity Analysis**

We conducted a sensitivity analysis that incorporated beneficiaries with dual diagnoses of both ILD and COPD into the COPD cohort.

**7A. Demographic and clinical characteristics of unique individuals at time of first year in the dataset (2016-2020)**

| | ILD (n=966) | COPD (n=73,743) | p-value[1] |
|---|---|---|---|
| Age, mean (SD) | 67.5 (15.4) | 64.6 (12.4) | <0.001 |
| Male, N (%) | 339 (35.1) | 30,136 (40.9) | <0.001 |
| Race/Ethnicity, N (%) | | | <0.001 |
| Non-Hispanic White | 545 (56.4) | 52,425 (71.1) | |
| Non-Hispanic Black | 177 (18.3) | 13,469 (18.3) | |
| Hispanic | 116 (12.0) | 3,775 (5.1) | |
| Other | 128 (13.3) | 4,074 (5.5) | |
| Long-term services and support use | | | 0.001 |
| Home and community-based services | 292 (30.2) | 18,711 (25.4) | |
| Months with an oxygen claim, median (IQR) | 12 (6-12) | 12 (6-12) | 0.397 |
| Non-metro resident, N (%) | 127 (13.2) | 14,202 (19.3) | <0.001 |
| Competitive bidding area resident, N (%) | 621 (64.3) | 43,414 (58.9) | 0.001 |
| Confounders of oxygen use, N (%) | | | |
| Prior or current tobacco use | 140 (14.5) | 30,695 (41.6) | <0.001 |
| Heart failure | 331 (34.3) | 21,908 (29.7) | 0.002 |
| Pulmonary hypertension | 134 (13.9) | 5,849 (7.9) | <0.001 |
| Count of other comorbidities, N (%) | | | 0.001 |
| 0 to 1 | 102 (10.6) | 8,305 (11.3) | |
| 2 to 3 | 187 (19.4) | 17,538 (23.8) | |
| 4 to 5 | 241 (25.0) | 19,476 (26.4) | |
| 6 to 7 | 234 (24.2) | 15,536 (21.1) | |
| 8 to 9 | 143 (14.8) | 9,133 (12.4) | |
| 10 or more | 59 (6.1) | 3,755 (5.1) | |
| Any Oxygen Claim, N (%) | 186 (19.3) | 13,906 (18.9) | 0.754 |
| High Flow (>4 L/min) Oxygen Claim, N (%) | --- | 358 (0.49) | -- |

[1] t-test for continuous variables; $chi^2$ test for categorical variables, rank sum test for months with oxygen claim

*categories with <10 observations are redacted

**7B. Results of a GEE model for repeated measures assessing factors influencing oxygen payments among beneficiaries who had claims paid for stationary oxygen concentrators**

| Variable | Coefficient | 95% CI | p-value |
|---|---|---|---|
| Age | -45.58 | -54.57, -36.60 | <0.001 |
| Male | 7.37 | -10.48, 25.22 | 0.418 |
| Race/ethnicity[1] | | | |
| *Non-Hispanic black* | 26.20 | 0.52, 51.88 | 0.046 |
| *Hispanic* | -12.71 | -60.27, 34.85 | 0.601 |
| *Other* | 51.24 | 8.58, 93.91 | 0.019 |
| Non-metro | 153.36 | 129.88, 176.84 | <0.001 |
| CBA | -65.53 | -86.48, -44.58 | <0.001 |
| Home and community-based services[2] | -93.75 | -11.02, -76.48 | <0.001 |
| Comorbidity count[3] | | | |
| *2-3* | -100.67 | -138.17, -63.17 | <0.001 |
| *4-5* | -151.11 | -187.73, -114.50 | <0.001 |
| *6-7* | -213.56 | -250.51, -176.60 | <0.001 |
| *8-9* | -286.74 | -325.21, -248.28 | <0.001 |
| *10 or more* | -326.34 | -368.86, -283.83 | <0.001 |
| Current or prior tobacco use | -49.17 | -67.24, -31.10 | <0.001 |
| Heart failure | 44.38 | 25.85, 62.91 | <0.001 |
| Pulmonary hypertension | 9.60 | -12.39, 31.60 | 0.392 |
| Gas oxygen (portable or stationary) | 176.65 | 159.40, 193.90 | <0.001 |
| Liquid oxygen (portable or stationary) | 202.68 | 72.33, 333.03 | 0.002 |
| Portable oxygen concentrator | 318.52 | 292.50, 344.55 | <0.001 |
| High flow (>4 L/min) | 29.40 | -17.44, 76.25 | 0.219 |
| ILD diagnosis | -6.80 | -97.79, 84.20 | 0.884 |
| _cons | 984.26 | 946.11, 1022.40 | <0.001 |

[1]Reference group = non-Hispanic white

[2]Reference group = 0-1 comorbidity

[3]Reference group = No home or community-based services

**7C. Estimated annual costs of oxygen equipment based on GEE model**

| Oxygen Equipment | ILD Annual Cost (95% CI) | COPD Annual Cost (95% CI) |
|---|---|---|
| Stationary concentrator only | $939.37 ($848.77, $1029.96) | $946.16 ($937.98, $954.35) |
| Stationary concentrator + Gas (Portable/Stationary) | $1002.42 ($911.67, $1093.17) | $1009.21 ($998.60, $1019.82) |
| Stationary concentrator + Liquid (Portable/Stationary) | $1141.12 ($985.82, $1296.42) | $1147.92 ($1017.76, $1278.07) |
| Stationary concentrator + Portable Concentrator | $1216.84 ($1123.83, $1309.86) | $1223.64 ($1199.07, $1248.21) |
